# Predicting Alcohol Dependence from Multi-Site Brain Structural Measures

**DOI:** 10.1101/2020.01.17.20016873

**Authors:** Sage Hahn, Scott Mackey, Janna Cousijn, John J. Foxe, Robert Hester, Kent Hutchinson, Ozlem Korucuoglu, Edythe London, Valentina Lorenzetti, Maartje Luijten, Reza Momenan, Catherine Orr, Martin Paulus, Lianne Schmaal, Rajita Sinha, Zsuzsika Sjoerds, Dan J. Stein, Elliot Stein, Ruth J. van Holst, Dick Veltman, Reinout W. Wiers, Murat Yucel, Paul M. Thompson, Patricia Conrod, Nicholas Allgaier, Hugh Garavan

**Author notes:** Correspondence: Sage Hahn, UVM Medical Center 1 South Prospect Street, Burlington Vermont 05401.

## Abstract

**Background:** The search for neuroimaging biomarkers of alcohol use disorder (AUD) has primarily been restricted to significance testing in small datasets of low diversity. To identify neurobiological markers beyond individual differences, it may be useful to develop classification models for AUD. The ever-increasing quantity of neuroimaging data demands methods that are robust to the complexities of multi-site designs and are generalizable to data from new scanners.

**Methods:** This study represents a mega-analysis of previously published datasets from 2,034 AUD and comparison participants spanning 27 sites, coordinated by the ENIGMA Addiction Working Group. Data were grouped into a training set including 1,652 participants (692 AUD, 24 sites), and test set with 382 participants (146 AUD, 3 sites). A battery of machine learning classifiers was evaluated using repeated random cross-validation (CV) and leave-site-out CV. Area under the receiver operating characteristic curve (AUC) was our base metric of performance.

**Results:** Multi-objective evolutionary search was conducted to identify sparse, generalizable, and high performing subsets of brain measurements. Cortical thickness in the left superior frontal gyrus and right lateral orbitofrontal cortex, cortical surface area in the right transverse temporal gyrus, and left putamen volume, appeared most frequently across searches. Restricting a regularized logistic regression model to these four features yielded a test-set AUC of .768.

**Conclusions:** Developing classification models on multi-site data with varied underlying class distributions poses unique challenges. Supplementing datasets with controls from new sites and performing feature selection increases generalizability. Four features identified by evolutionary search may serve as specific biomarkers for individuals with current AUD.

## Introduction

While the evidence associating alcohol use disorder (AUD) with structural brain differences is strong, there is considerable merit in establishing robust and generalizable neuroimaging-based AUD biomarkers. These biomarkers would have objective utility for diagnosis and may ultimately help in identifying youth at risk for AUD and for tracking recovery and treatment efficacy in abstinence, including relapse potential. While these types of clinical applications have not yet been realized with neuroimaging, current diagnostic practices are far from perfect: The inter-observer reliability of past year AUD, as diagnosed by the DSM-IV, was calculated with Cohen’s kappa as .74±.09 (1). More immediately, neurobiological markers of AUD can give clues to potential etiological mechanisms.

Here, we apply a supervised learning approach, in which a function is trained to map brain structural measures to AUD diagnosis, and then evaluated on unseen data. Prior approaches to developing machine learning classifiers for AUD include a similar binary machine learning classification approach discriminating between AUD and substance naive controls (2). Their analysis made use of 296 participants, case and control, and reported a leave-one-out cross validated (CV) balanced accuracy of 74%. A further example of recent work includes that by Adeli et al. on distinguishing AUD from controls (among other phenotypes), on a larger sample of 421, yielding a balanced accuracy across 10-fold CV of 70.1% (3). In both examples, volumetric brain measures were extracted and used to train and evaluate proposed machine learning (ML) algorithms. The present study differs from prior work in both its sample size (n=2,034) and complex case to control distribution across many sites. Mackey et al. developed a support vector machine (SVM) classifier that obtained an average area under the receiver characteristic operator curve (AUC) of .76 on a subset of the training data presented within this work (4). Our present study expands on this previous work by exploring an extensive range of classifiers using new, additional samples. Further, we explore the importance of the specific cross validation schemes that can be employed, address matters of residualizing data for known covariates, and also focus on an approach to determine robust feature importance.

An important consideration for any large multi-site neuroimaging study, particularly relevant in developing classifiers, is properly handling data from multiple sites (5). Any useful classifier should generalize to new data, possibly from a different scanner or country. In this sense, any information gleaned from a classifier that generalizes poorly to unseen data is unlikely to represent the actual effect of interest. More generally and within the broader field of ML, the task of creating “fair” or otherwise unbiased classifiers has received a great deal of attention (6). In our study, the imbalance between numbers of cases and controls across different sites is a significant challenge, as unrelated, coincidental scanner or site effects may easily be exploited by multivariate classifiers - leading to spurious or misleading results.

A related consideration is how one should interpret the neurobiological significance of features that contribute most to a successful classifier. We propose a multi-objective genetic algorithm (GA) based feature selection search to isolate meaningful brain measures and tackle the complexities of handling differing class distributions across sites. GA are considered a subset of evolutionary search algorithms within the broader field of artificial intelligence. A sizeable body of research has been conducted into the usage of multi-objective genetic algorithms, introducing a number of effective and general techniques to navigate high dimensional search spaces, e.g., various optimization and mutation strategies. (7,8,9). Our proposed GA is designed to select a set of features both useful for predicting AUD and generalizable to new sites. By selecting not just predictable, but explicitly generalizable and predictable features, we hope to identify features with true neurobiological relevance. We draw motivation from a large body of existing work that has successfully applied GAs to feature selection in varied machine learning contexts (10, 11).

This study represents a continuation of work by Mackey et al. and the Enhancing Neuro-Imaging Genetics through Meta-Analysis (ENIGMA) Addiction Working Group (http://enigmaaddiction.com), in which neuroimaging data were collected and pooled across multiple laboratories to investigate dependence on multiple substances (4). Here, we focus on a more exhaustive exploration of machine learning to distinguish AUD from non-dependent individuals, spanning 27 different sites. We evaluate the role of varied cross-validation (CV) strategies in addition to the optional inclusion of control-only sites. We also introduce methods suitable for complex multi-site data with varied underlying class distributions. Finally, we present classification results for a left-out testing set sourced from three unseen sites, as a measure of classifier generalizability.

## Methods and Materials

### Dataset

Informed consent was obtained from all participants and data collection was performed in compliance with the Declaration of Helsinki. Individuals were excluded if they had a lifetime history of any neurological disease, a current DSM-IV axis I diagnosis other than depressive and anxiety disorders, or any contraindication for MRI. A variety of diagnostic instruments were used to assess alcohol dependence (4).

Participants’ structural T1 weighted brain MRI scans were first analyzed using FreeSurfer 5.3 which automatically segments 14 bilateral subcortical regions of interest (ROIs) and parcellates the cortex into 78 bilateral ROIs, for a total of 150 different measurements representing cortical mean thickness and surface area along with subcortical volume according to the Desikan parcellation (12, 13). Quality control of the FreeSurfer output including visual inspection of the ROI parcellations was performed at each site according to the ENIGMA protocols for multi-site studies, available at: http://enigma.ini.usc.edu/protocols/imaging-protocols/. In addition, a random sample from each site was examined at a central location to ensure consistent quality control across sites. All individuals with missing volumetric or surface ROIs were excluded from analyses.

In total, 2,034 participants from 27 different sites met all inclusion criteria. Further, data were separated into a training set composed of 1,652 participants (692 with AUD), from 24 sites with the remaining 382 participants (146 with AUD) from 3 sites isolated as a test set. The testing set represents a collection of new data submitted to the consortium that was not included in the most recent working group publication (4), constituting as realistic a test set as possible. **Table 1** presents basic demographic information on training and test splits. Within the training set, two sites contained only cases, 14 sites included only controls, and five sites contained an unbalanced mix in the number of cases and controls. **Figure** 1 shows the distribution by site, broken down by AUD versus control.

**Table 1:**
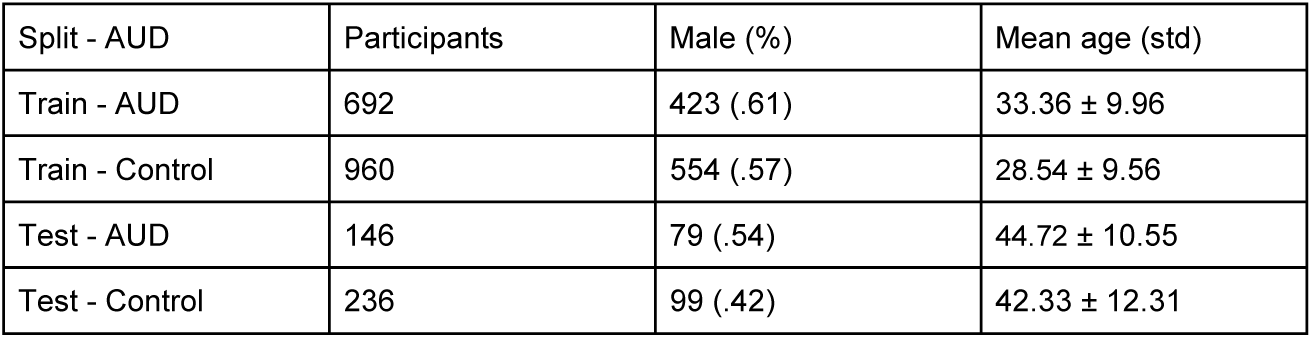
Sex and Age, across the full collected dataset from 27 sites as split further into training and withheld testing set, and by alcohol use disorder (AUD) vs. control.

### Exploratory Data Analysis

In this section, we describe an exploratory analysis investigating different choices of training data, classification algorithm, and cross-validation strategy. A final framework for training is determined from this exploration, and its implementation and evaluation are covered in the following sections.

We explored classifier performance first on a base training dataset (**Figure 1**, sites 1-8), composed of only sites containing at least one case participant. The same experimental evaluation was then repeated with an augmented version of the dataset, adding additional participants from 16 control-only sites (**Figure 1**, sites 9-24). The top row of **Figure 2** outlines these two options within the context of our experimental design.

**Figure 1:**
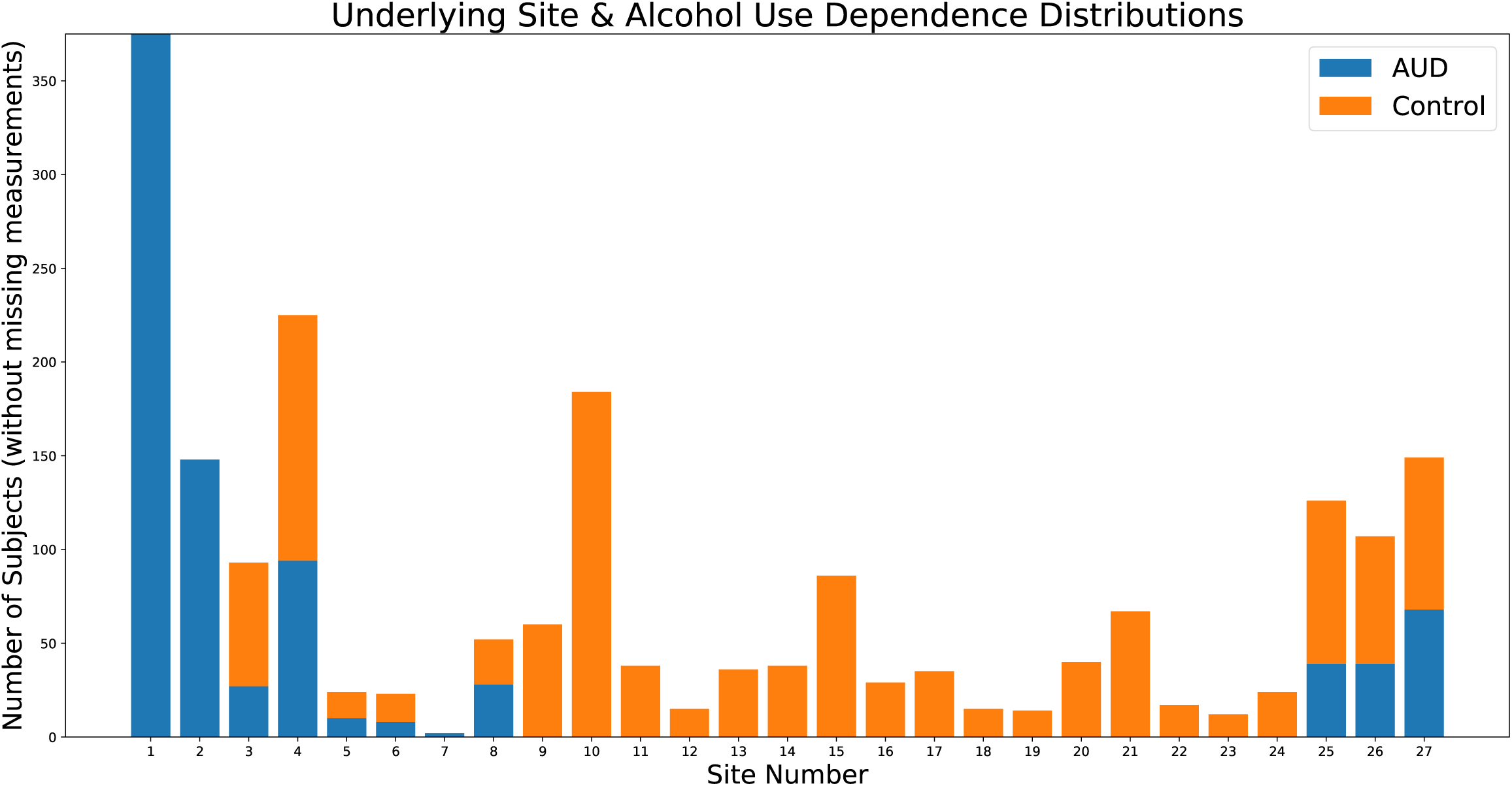
The distribution of both training (Sites 0-23) and testing (24-26) datasets is shown, and further broken down by AUD to case ratio per site.

**Figure 2:**
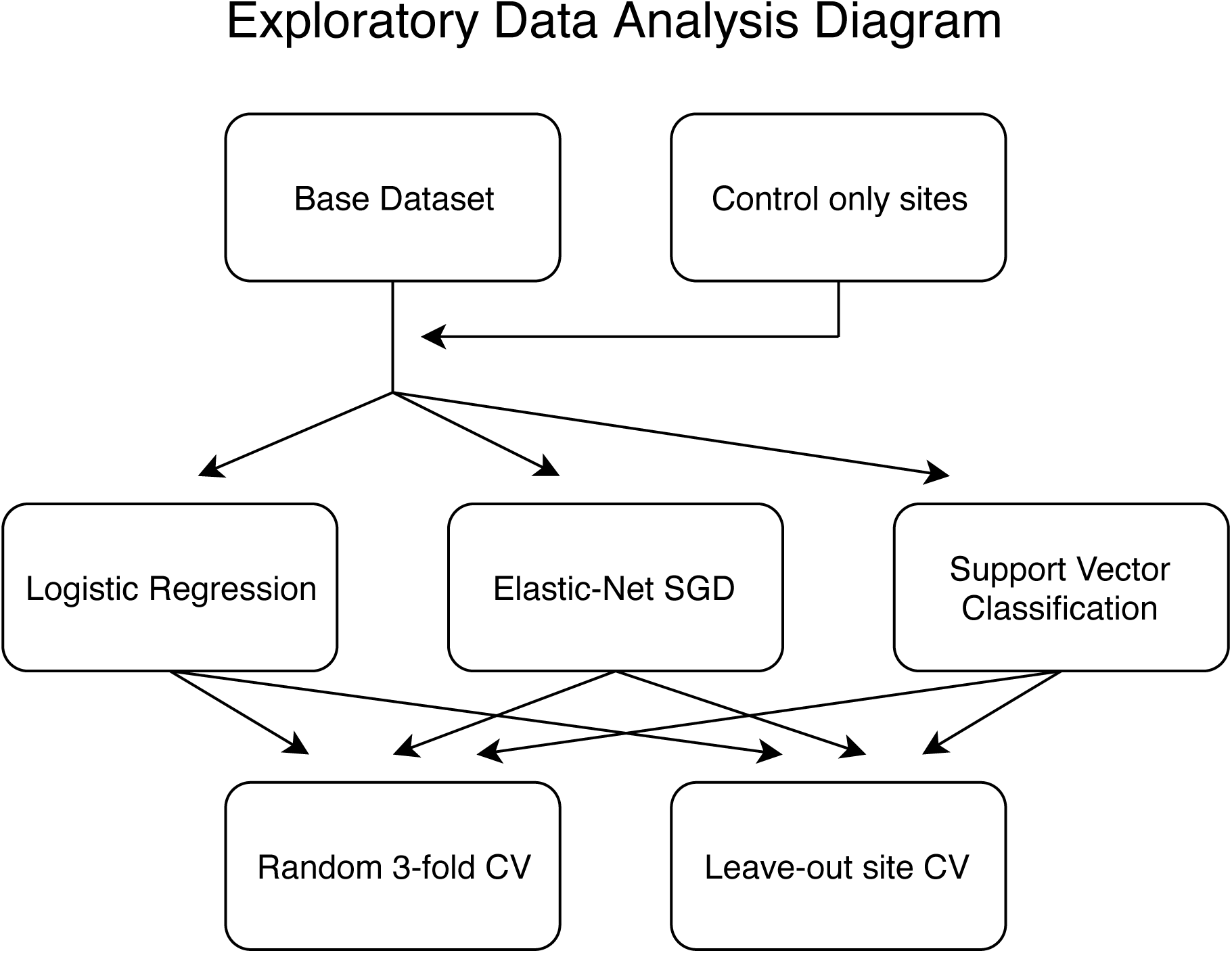
The different permutations of analyses conducted internally on the training set, with differing input dataset options (top row), classifiers (middle row) and computed CV scoring metrics (bottom row).

Three machine learning algorithms suitable for binary classification (**Figure 2**, middle row) were implemented within the python library Scikit-learn (14). Most simply, we considered a regularized ridge logistic regression classifier (l2 loss) with regularization parameter values chosen through an internal CV. Another variant of regularized logistic regression optimized with stochastic gradient descent (SGD) was implemented with an elastic net loss (l1 and l2). A nested random parameter search was conducted, across 100 values, determining the choice of loss function and regularization values (15). Finally, we considered a Support Vector Machine (SVM) with radial basis function (rbf) kernel, which allowed the classifier to learn nonlinear interactions between features (16). Like the hyperparameter optimization strategy employed for the SGD logistic regression, a random search over 100 SVM parameter combinations, with differing values for the regularization and kernel coefficients, was employed with nested CV for parameter selection. Exact training details are provided in the supplemental materials.

Proper CV is of the utmost importance in machine learning applications. It is well known that - if improperly cross-validated - classifiers can overfit onto validation sets, and even with more sophisticated CV techniques can overestimate expected generalization (17). Within this work, we made use of standard random repeated 3-fold CV, where an indication of classifier performance is given by its averaged performance when trained on one portion of the data and tested on a left-out portion, across different random partitions (18). We also made use of a leave-site-out CV scheme across the five sites that include both cases and controls. These options are shown in the bottom row in **Figure 2**. We computed metrics according to both schemes for all considered classifiers on the training dataset. Area under the Receiver Operating Characteristic curve (AUC) was used as a base performance metric insensitive to class imbalance (19).

### Final Analytic Pipeline

We implemented a GA designed to select sets of features most useful in training a site generalizable classifier. Towards this goal, the GA repeatedly trained and evaluated a regularized logistic regression classifier, as introduced earlier, on initially random subsets of possible features. These feature subsets were then optimized for high AUC scores as determined by the leave-out site CV using the five sites that include both cases and controls. Multi-objective optimization was conducted with the aid of a number of successful GA strategies, and these include: random tournament selection (20), feature set mutations, repeated runs with isolated populations, a sparsity objective similar in function to “Age-fitness Pareto optimization” (21), among others. An introduction to GA and a complete description of our design decisions regarding the algorithm are provided in the supplemental material.

The algorithm was run across six different variants of hyperparameters, as shown in **Figure 3.** We explored choices related to size (number of subsets of features considered in each round) and scope (how many times the search is iterated) in addition to objective functions. The results from each search variant represent thousands of subsets of features, each with an associated performance score. We restricted the output from each search to only the top 200 - and therefore to high performing - feature subsets. The final feature subsets were ultimately pooled together and considered in a feature importance meta-analysis. In determining feature importance, the following considerations were used: each subset’s individual performance (higher performance weighted higher) and the number of features (subsets with more features were penalized). A final measure of feature importance was calculated as the average feature importance from each of the six search variants computed separately. Within each search variant, an individual feature’s importance was defined as the sum of a feature set’s fitness scores, further divided by the number of total features in that set, across all the top 200 sets in which that feature appeared. Importances per set were then normalized, such that intuitively a feature present in all of the top 200 feature sets would have a value of 1, and if present in none, 0. Each feature’s final score therefore represents that feature’s averaged score (between 0 and 1) as derived from each separate search. We were interested at this stage in identifying a relative ranking of brain features, as, intuitively, some features should be more helpful in classifying AUD, and features that are useful towards classification are candidates to be related to the underlying AUD neurobiology.

**Figure 3:**
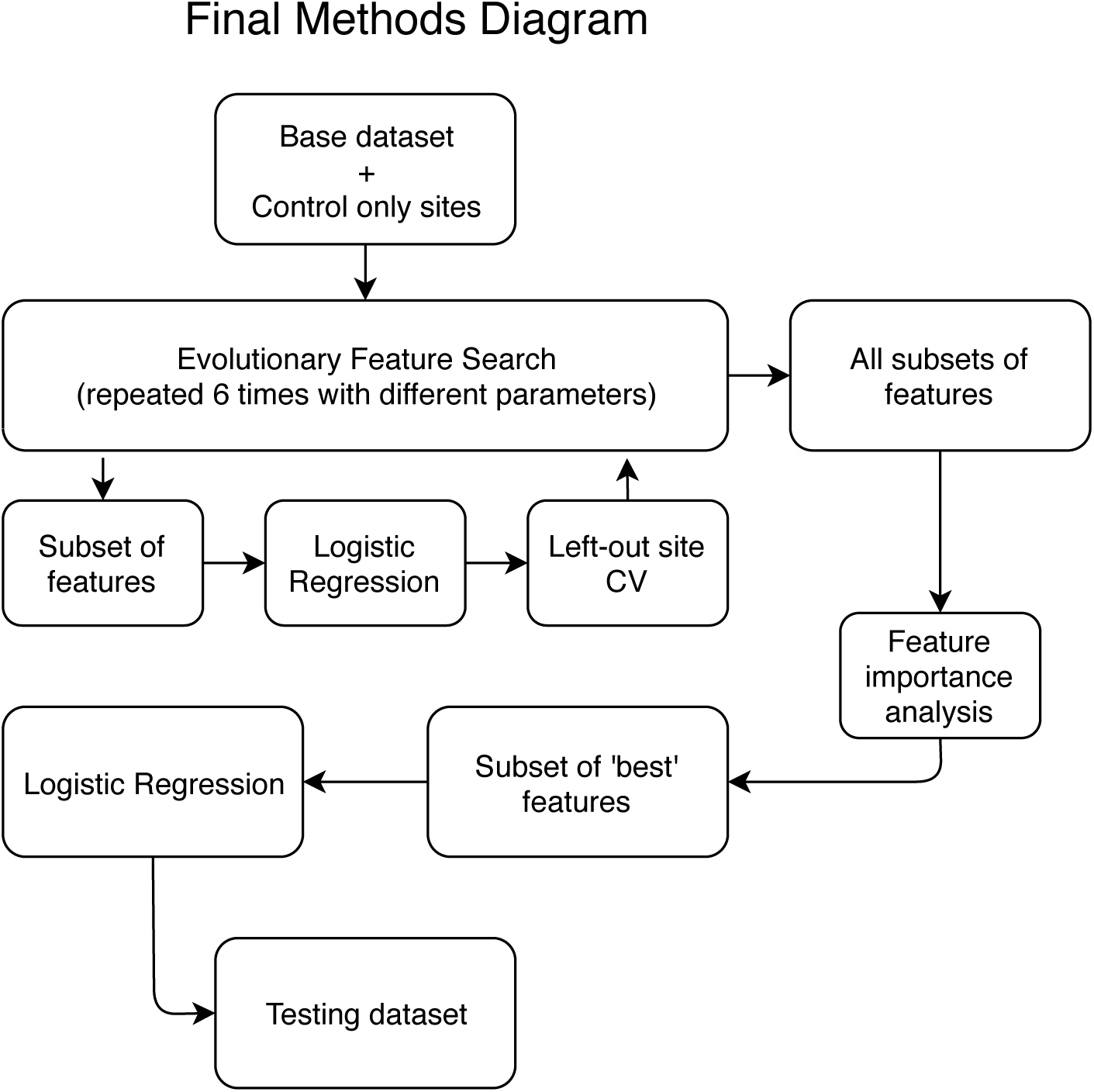
A simplified view of the final pipeline, where the full training dataset is employed in an evolutionary feature search designed to produce optimal subsets of high performing features. From this collection of feature subsets a meta-analysis for determining feature importance is conducted and a subset of ‘best’ features are selected. Next, a logistic regression classifier is trained and evaluated on the testing dataset, with access to only the ‘best’ subset of features.

As referenced in **Figure 3**, we selected a “best” subset of features with which to train and evaluate a final regularized logistic regression classifier on the withheld testing set. We determined the “best” subset of features to be those which obtained a final feature importance score above a user defined threshold. Ideally, this threshold would be determined analytically on an additional independent validation sample by relating classifier performance, with access to only those features over threshold, to that threshold. With limited access to data from case-control balanced sites, we employed only internal CV towards the choice of threshold. Post-hoc analyses were conducted with differing thresholds, providing an estimate as to how important this step may prove in future analyses.

## Results

### Exploratory Data Analysis

The complete exploratory training set results are shown in **Table 2**. Regularized logistic regression on the base dataset yielded an AUC of .907 ± .022 (standard deviation across folds) under 3-Fold CV versus .560 ± .189 under leave-out-site, and with added controls an AUC of .917 ± .010 with random CV and .636 ± .169 with leave-out-site. The choice of classifier produced only minor differences in performance (±.02), regardless of the CV method. Including additional control participants yielded a small boost to random 3-fold CV scores (.003 - .023), and a more noticeable gain to leave-out site CV scores (.053 - .091). The CV strategy (Random vs. Leave-out-site) produced the largest discrepancy in scores (.267 - .347) with the former yielding inflated results.

**Table 2:** The results for each of the three considered classifiers with and without extra control participants (see **Figure** 1 for sites deemed extra control, namely those containing only control participants) across both cross validation (CV) strategies, as highlighted in **Figure** 2. Standard deviation in area under the receiver characteristic operator curve (AUC) across cross-validated folds is provided, as an estimate of confidence.

| Dataset | Classifier | Random 3-Fold CV AUC | Leave-out 5 Site CV AUC |
| --- | --- | --- | --- |
| Base | Logistic Regression | .907 ± .022 | .560 ± .189 |
| Base | SGD | .896 ± .012 | .561 ± .183 |
| Base | SVM | .912 ± .011 | .578 ± .111 |
| Base + Control only | Logistic Regression | .917 ± .012 | .636 ± .169 |
| Base + Control only | SGD | .919 ± .009 | .652 ± .132 |
| Base + Control only | SVM | .915 ± .014 | .631 ± .139 |

### Feature Importance

The top 15 features as determined by average weighted feature importance, from all six searches (i.e., base training dataset only and base plus control-only datasets, by three machine-learning algorithms; see **Figure 2**), are presented in **Figure 4**. Four features emerged with an importance score greater than .8 (where an importance score of 1 represents a feature present in every top feature set and 0 in none), followed by a slightly sharper decline and, not shown, a continuing decline. Also shown are the cortical surface area and thickness features as projected onto the *fsaverage* cortical surface space. The left putamen (.816) and left pallidum (.210) volumes were the only subcortical features with feature importance scores over .05 (not shown).

**Figure 4:**
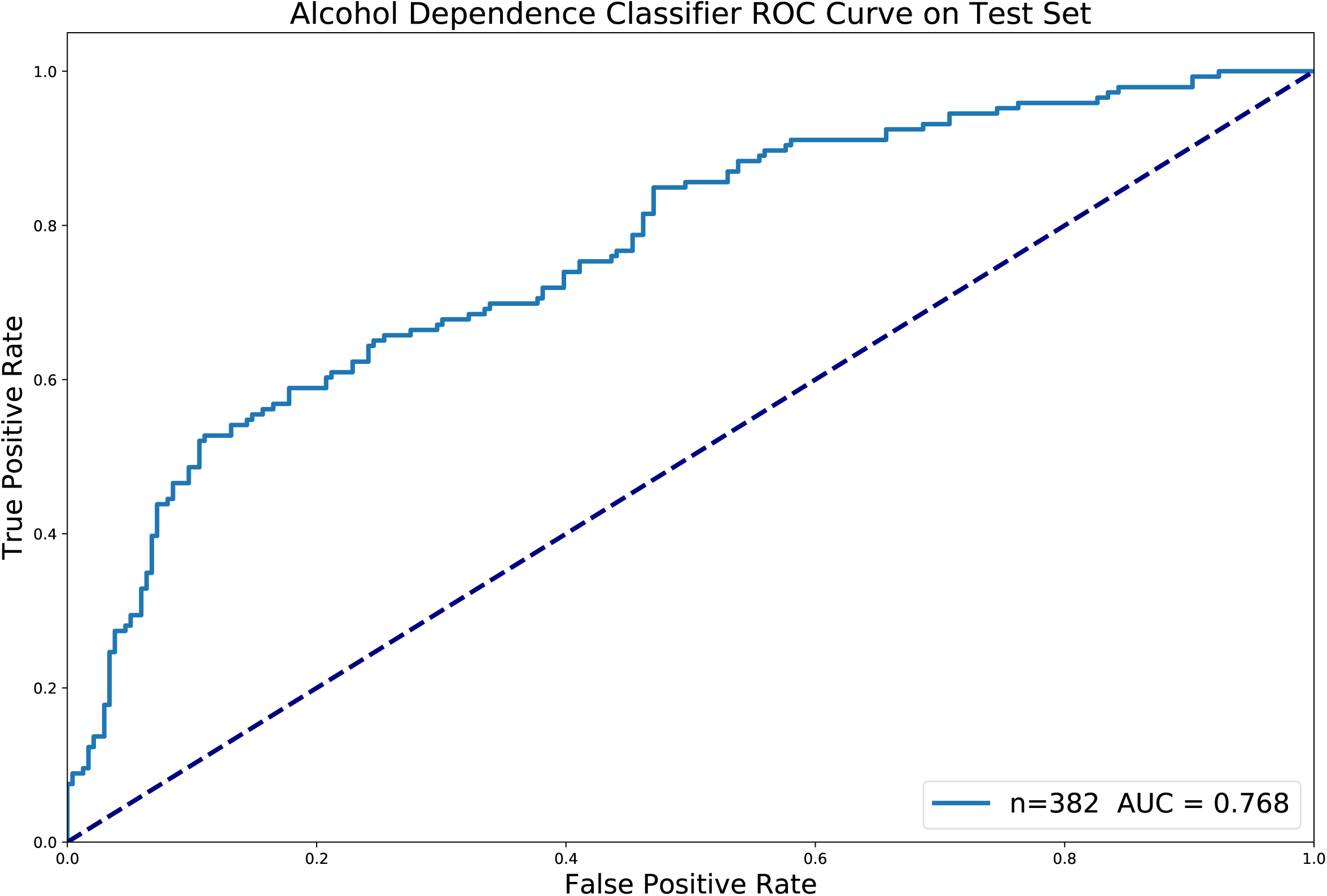
(A) The top 15 features, as ranked by average weighted feature importance (where 0 indicates a feature appeared in none of the GA final models, and 1 represents a feature appeared in all) are shown. (B) The cortical thickness and (C) cortical average surface area feature importance scores, above a threshold of .1, are shown as projected onto the fsaverage surface space.

### Testing Set Evaluation

Further internal nested validation on the training selected a threshold of .8 weighted feature importance and above, which corresponds to the top four features only (**Figure 4**). The final model, trained on only this “best” subset of four features, achieved an AUC of .768 on the testing set. The ROC curve for this classifier on the testing set is shown in **Figure 5**. We further conducted a number of post-hoc analyses on the testing dataset. To confirm the predictive utility of GA feature selection, a regularized logistic regression model and SVM model with access to all features were both trained on the full training dataset and evaluated on the testing set. The logistic regression scored .697 AUC and the SVM .673 AUC. Similarly, regularized logistic regression and SVM models were trained on all features, but without the inclusion of additional control-only sites, and scored respectively .647 and .609 AUC. The final model was better than both the logistic regression model with all features and subjects (p=.0098) and without control subjects (p=3.5×10^−5^). We further investigated the choice of user defined threshold in selecting number of top features by testing the inclusion of top 2 to 15 features. Some notable differences can be seen in performance, for example: .782 AUC with top three, .737 AUC with top five and .741 AUC with top ten.

**Figure 5:**
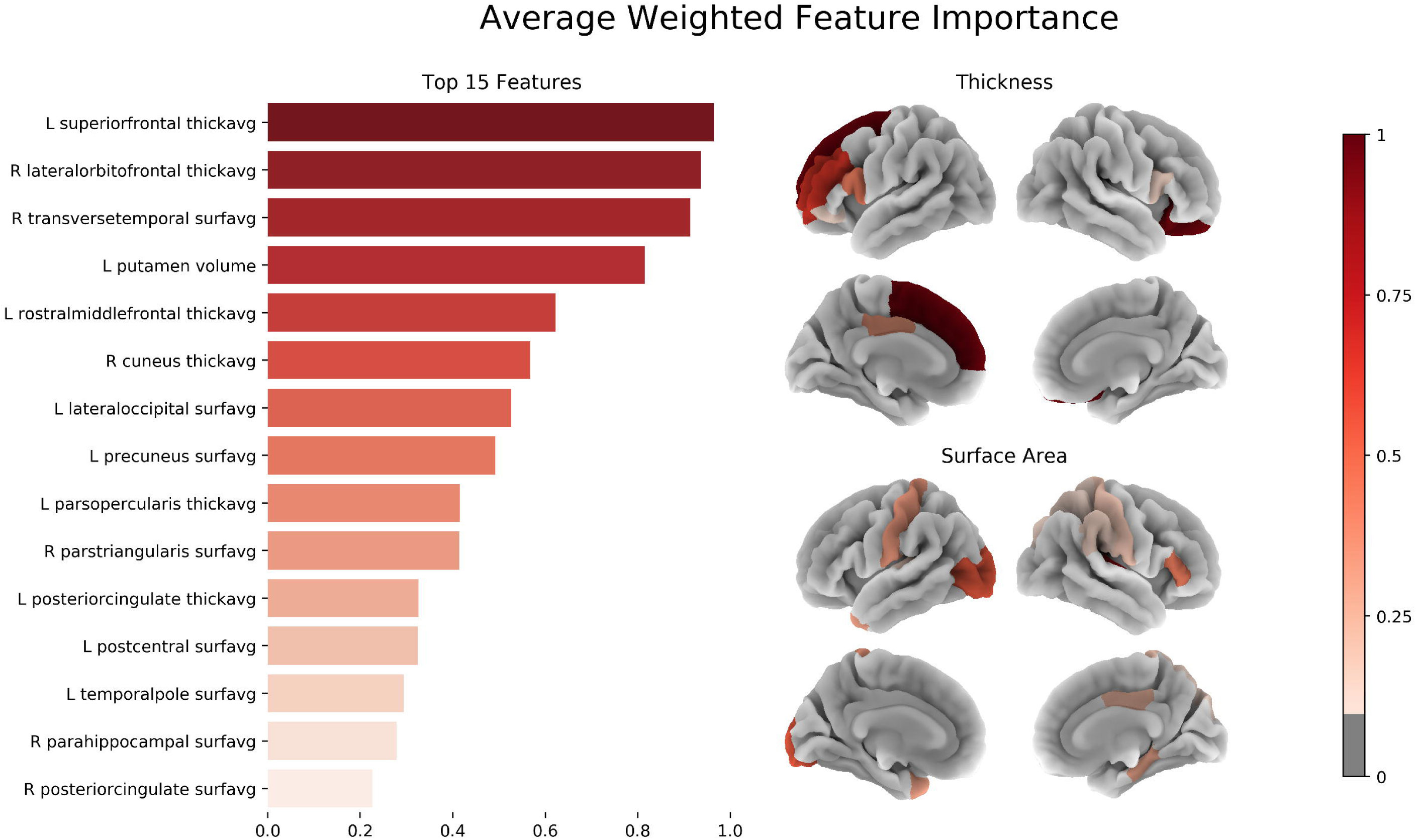
The ROC curve for the final logistic regression model on the testing set, as restricted to only the “best” subset of 4 features.

## Discussion

We used multi-site neuroimaging data to identify structural brain features that classify new participants, from new sites, as having an AUD with high accuracy. In doing so, we highlighted the importance of carefully chosen metrics in accurately estimating ML classifier performance in the context of multi-site imbalanced neuroimaging datasets. We further explored several techniques, ranging from analytical methods to more general approaches, and their merit towards improving classifier performance and generalizability. Our proposed GA-derived feature importance measure, in addition to aiding classification, might help in identifying neurobiologically meaningful effects.

A clear discrepancy arose between random repeated CV (i.e., participants randomly selected from across sites) and leave-site-out CV results (**Table 2**). We suspect that the random repeated CV overestimates performance due to covert site effects. The classifiers appeared to memorize some set of attributes, unrelated to AUD, within the case- and control-only sites, and therefore were able to accurately predict AUD only if participants from a given site were present in both training and validation folds. Performance on leave-site-out CV, in contrast to random repeated CV, better estimates classifier generalizability to new unseen sites. This is validated by post hoc analyses in which a logistic regression trained on all features obtained a test set AUC (.697) far closer to its training set leave-out site CV score (.636 ± .119) then its random repeated CV score (.917). While this observation must be interpreted within the scope of our presented imbalanced dataset, these results stress the importance of choosing an appropriate performance metric, and further highlight the magnitude of error that can be introduced when this metric is chosen incorrectly.

In addition to performing model and parameter selection based on a more accurate internal metric, the addition of control-only participants proved beneficial to classifier performance (.053 - .091 gain in leave-out site AUC). This effect can be noted within our exploratory data analysis results (**Table 2**) comparing leave-site-out CV results between the base-only dataset and the base plus control-only site dataset. When extra control participants are added performance increased up to .09 AUC. Post-hoc analysis revealed a similar performance gain on the testing set from adding control participants; logistic regression plus .05 AUC and SVM plus .06 AUC. This boost likely reflects a combination of two circumstances. In the first, the underlying ML algorithm is aided by both more data points to learn from and a more balanced case to control distribution, which have both been shown to aid binary classification performance (22). The second reflects a resistance to the learning of site-related effects which, as noted above, can lead to the algorithm detrimentally learning covert site effects. This is potentially due to the inclusion of more sites and scanners, which makes the unintentional learning of specific site effects (as a proxy for AUD) more difficult. As neuroimaging data banks continue to grow, the potential arises for supplementing analyses with seemingly unrelated external datasets in clever and powerful ways.

Our proposed GA-based feature selection, with inclusion of leave-site-out criteria, proved to be useful in improving classifier generalizability. This is highlighted by a .071 boost to AUC score in a model trained on only the top identified four features in contrast to a model trained with all the available features. We believe the observed performance boost to be a result of only allowing the classifier to learn from features previously determined to be useful towards site generalizable classification. In this way, the final classifier can avoid adverse site effects through a lack of exposure to brain measurements highly linked to specific sites. Further post-hoc results indicate even higher performance with just the top three features (+.014 vs. selected top four feature model), and a slight decrease with the addition of more features. While these results are post-hoc, they do suggest an inherent benefit to sparsity of selected features. This falls in line with the general understanding of the generalizability versus overfitting tradeoff, where limiting the number of input features can function as regularization, in this case limiting a model’s ability to overfit by not providing it access to spurious features. In future work, an additional validation set might prove useful in selecting between different final models and thresholds, in addition to careful comparisons between different feature selection methods.

A persistent issue in typical interpretation from ML models is the issue of shared variance between different features. The features a single model selects may very well have suitable surrogate features within the remaining dataset. In contrast, our feature importance metric is derived from thousands of models, providing the chance for equivalent features, with shared variance, to achieve similar importance scores. A natural distinction nevertheless exists between predictive features and those that emerge from univariate testing as significant. Specifically, the absence of a feature within our final model, (i.e., unimportance of a feature by our metric), does not necessarily imply a lack of association between that feature and AUD. An absence could alternatively indicate that a different feature better captures some overlapping predictive utility, which is different conceptually from sharing variance in that in this case one feature is consistently more useful for prediction. The redundant feature therefore might not appear as important despite an association with AUD when considered in isolation. On the other hand, a feature with a relatively weak association could emerge with consistently high feature importance if it proves uniquely beneficial to prediction. Above and beyond univariate significance, if a given feature does have predictive utility, it strongly suggests that a real association exists. Our selected top features were both identified as consistently useful features within the training set and experimentally confirmed as site generalizable on the testing set.

The top four features as identified by our introduced metric of feature importance were the average cortical thickness of the left superior frontal gyrus and right lateral orbitofrontal cortex (OFC), the left putamen volume and the average surface area of the right transverse temporal gyrus. Specifically, cortical thinning, volume and surface area reduction across these regions prompt the trained model towards an AUD prediction. Thinning, within the left superior frontal gyrus and right lateral OFC, agrees broadly with the literature which has consistently shown frontal lobe regions to be most vulnerable to alcohol consequences (23). Prefrontal cortical thinning and reduced volume in the left putamen seem to further indicate specific involvement of the mesocorticolimbic system. This dopaminergic brain pathway has been consistently linked with alcohol dependence and addiction in general (24, 25). Likewise, a recent voxel-based meta-analysis showed a significant association between lifetime alcohol consumption and decreased volume in left putamen and left middle frontal gyrus (26).

Comparing the four selected regions in the present study with those determined to be significant by univariate testing on an overlapping dataset from Mackey et al. 2019, we find three regions in common (the exception being right transverse temporal gyrus surface, as surface area was not considered in that analysis). Further, left superior frontal and putamen appeared as two of the top 20 features in both folds of an SVM classifier trained and tested on split halves in the Mackey paper (right lateral orbital frontal only appeared in one-fold). Of the existing alcohol classifiers mentioned in the introduction by Guggenmos et al. (2018) and Adeli et al. (2019), only Adeli reported overlapping AUD associated regions with our top four: lateral orbitofrontal thickness and superior frontal volume.

In interpreting the performance of a classifier linking brain measurements to an external phenotype of interest, we also need to consider how reliably the phenotype can be measured. The exact relationship between interobserver variability of a phenotype or specific diagnosis and ease of predictability or upper bound of predictability is unknown, but it seems plausible that they would be related. This proves pertinent in any case where the presented ground truth labels, those used to generate performance metrics, are noisy. We believe further study quantifying these relationships will be an important next step towards interpreting the results of neuroimaging-based classification, as even if a classifier capable of perfectly predicting between case and control existed, it would be bound by our current diagnostic standard. A potential route towards establishing a robust understanding of brain changes associated with AUD might involve some combination of standard diagnostic practices with objective measures or indices gleaned from brain-based classifiers. Relating classifiers directly with specific treatment outcomes (potential index for recovery), or within a longitudinal screening context (potential index for risk) represent further exciting and useful applications.

We have drawn attention to the impact on model generalizability of case distribution by site within large multi-site neuroimaging studies. In particular, we have shown that CV methods that do not properly account for site can drastically overestimate results, and presented a leave-site-out CV scheme as a better framework to estimate model generalization. We further presented an evolutionary-based feature selection method aimed at extracting usable information from case- and control-only sites, and showed how this method can produce more interpretable, generalizable and high-performing AUD classifiers. Finally, a measure of feature importance was used to determine relevant predictive features, and we discussed their potential contribution to our understanding of AUD neurobiology.

## Data Availability

Data is not publically available but accessed through enigma addiction data pooling.

## Acknowledgements

This work was made possible by NIDA grant R01DA047119 awarded to Dr. Garavan. Sage Hahn was supported by NIDA grant T32DA043593. Data collection was made possible through the following grants: Dr. Korucuoglu received support for the Neuro-ADAPT study from VICI grant 453.08.01 from the Netherlands Organization for Scientific Research (NWO), awarded to Reinout W. Wiers. Drs. Schmaal and Veltman received funding from Netherlands Organization for Health Research and Development (ZonMW) grant 31160003 from NWO. Drs. Sjoerds and Veltman received funding from ZonMW grant 31160004 from NWO. Dr. van Holst received funding from ZonMW grant 91676084 from NWO. Dr. Luijten and Veltman received funding from VIDI grant 016.08.322 from NWO, awarded to Dr. Cousijn received funding for the Cannabis Prospective study from ZonMW grant 31180002 from NWO. Drs. Garavan and Foxe received funds from NIDA grant R01-DA014100. Dr. London was supported by NIDA grant R01 DA020726, the Thomas P. and Katherine K. Pike Chair in Addiction Studies, the Endowment From the Marjorie Greene Family Trust, and UCLA contract 20063287 with Philip Morris USA. Data collection by Dr. Momenan was supported by the Intramural Clinical and Biological Research Program funding ZIA AA000125-04 DICB (Clinical NeuroImaging Research Core to RM) of the National Institute on Alcohol Abuse and Alcoholism (NIAAA). Dr. Paulus received funding from NIMH grant R01 DA018307. Dr. Stein was supported by the Intramural Research Program of NIDA and NIH. Dr. Sinha received funds from NIDA (PL30–1DA024859–01), the NIH National Center for Research Resources (UL1-RR24925–01), and NIAAA (R01-AA013892). Prof. Yücel was supported by National Health and Medical Research Council Fellowship 1117188 and the David Winston Turner Endowment Fund. Dr. Thompson was supported in part by NIH grant U54 EB020403.

## Disclosures

Dr. Sinha has served on the scientific advisory board of Embera Neuro-therapeutics. DJS has received research grants and/or consultancy honoraria from Lundbeck and Sun. Prof. Yücel has received funding from several law firms in relation to expert witness reports. PT received grant support from Biogen, Inc., for research unrelated to this study. All other authors have no financial disclosures.

